# Heart failure admission characteristics in women with a history of sexual or physical abuse: a case-control study

**DOI:** 10.1101/2024.03.26.24303298

**Authors:** Alicia Chan, Suzanne M. Cosh, John Horowitz, Phillip J. Tully

**Affiliations:** Department of Cardiology, Queen Elizabeth Hospital, Woodville, Australia; School of Psychology, The University of New England, Armidale, Australia; Cardiology Research Laboratory, Basil Hetzel Institute for Translational Health Research, Queen Elizabeth Hospital, Adelaide, Australia; School of Medicine, The University of Adelaide, Adelaide, Australia

**Keywords:** heart failure, intimate partner violence, child abuse, sexual assault

## Abstract

**Introduction:** An emerging body of epidemiological evidence links a history of exposure to sexual and physical abuse or assault with an increased risk of developing cardiovascular disease. Understanding adverse physical health outcomes including heart failure (HF) in persons exposed to sexual and physical abuse is of particular importance to help improve multidisciplinary approaches to mental and physical wellbeing, including cardiovascular management, among vulnerable populations.

**Methods:** This case-control study was performed at a tertiary hospital in metropolitan South Australia. At index HF admission 12 consecutive female patients with exposure to sexual and physical abuse or assault were identified from the trauma module of a structured psychiatric interview. Index admission data were classified into HF phenotype and aetiology using the criteria and definitions of the European Society of Cardiology. The presentation profile at index HF admission was compared to 12 gender and age-matched comparators.

**Results:** The clinical characteristics at index admission showed that most patients presented with pulmonary rales, peripheral oedema, and pulmonary congestion (67-75%), with diuretics the most common intervention (75%). By comparison to the non-exposed group, we observed differences between groups in HF severity determined by New York Heart Association class, diastolic blood pressure (BP), and heart rate (HR) but not systolic BP, saturation of peripheral oxygen (Sp02) or HF phenotypes. There were no differences in troponin, creatinine, sodium, admission use of inotropes, vasodilators or diuretics, post-traumatic stress disorder or comorbidities between the exposed patients and age-matched female controls.

**Conclusion:** Women with a history of experiencing sexual and physical abuse or assault presented with a unique HF admission profile characterised by lower BP and HR. The findings support previous work suggesting that persons experiencing sexual and physical abuse or assault have blunted cardiovascular reactivity to stressors such as hospital admission.

## INTRODUCTION

A solid body of empirical evidence links post-traumatic stress disorder (PTSD) with heart failure (HF) [1]. A parallel body of research links exposure to sexual abuse, violence, and intimate partner violence (IPV) with an increased risk of incident cardiovascular disease (CVD) [2, 3] as well as metabolic disease such as diabetes [4, 5]. Indeed, psychosocial stressors including PTSD, depression, and anxiety - all common comorbidities in survivors of abuse [6] - are also recognised as risk factors for CVD [7] in International Cardiology guidelines [8, 9]. Collectively, these findings raise the possibility that exposure to sexual abuse, violence, and intimate partner violence (IPV), with or without PTSD, are themselves risk factors for HF. However, to date, no previous research has described the association between such adverse and traumatic events with the presentation of HF in clinical practice. This is an important limitation to reconcile given the public health importance of HF worldwide [1, 10], coupled with the adverse mental health and cardiovascular outcomes for women exposed to sexual and physical abuse, assault or IPV [2, 3], and the importance of trauma-informed healthcare to optimise health outcomes in this population [11]. Accordingly, this study aims to characterise the HF admission characteristics, phenotypes, treatments, and outcomes of women exposed to sexual and physical abuse, assault or IPV.

## MATERIALS AND METHODS

The study was performed in the Central and Northern Adelaide Health Service, a metropolitan area in Adelaide, South Australia. We examined the medical records at index HF admission for twelve consecutive female patients with the exposure of interest and compared them to their age-matched and unexposed counterparts with HF. Index admission data were retrospectively classified, blind to patient exposure, into a single HF phenotype and aetiology by a single cardiologist (AC) using the criteria and definitions of the European Society of Cardiology [12]. Other data audited by the cardiologist to characterise the index HF admission included comorbidities, troponin T (TNT), creatinine, sodium, admission use of inotropes, vasodilators, and diuretics. The measurement of NTpro B-type natriuretic peptide and brain-derived neurotrophic factor is not routinely collected at admission in our health service. The exposure to trauma was determined by a two-stage psychosocial screening for depression and anxiety (PRIME-MD) [13] followed by a Structured Clinical Interview for common mental disorders (SCID-I, II) [14]. The trauma module of the structured interview was used to classify the exposure into a single variable, defined as any physical abuse, family violence, or IPV at any age (n=4), sexual abuse or sexual assault at any age (n=5), both a physical and sexual abuse (n=3).

Patients provided written consent and this study complies with the Declaration of Helsinki (Human Research Ethics Committee approval from the Queen Elizabeth and Lyell McEwin hospitals TQEHLMH/188). Due to the potential for re-identifying sensitive information about the exposure of interest, only general descriptive information is provided for each case. Female patients with the exposure of interest were matched by age (1:1 ratio) to other female HF patients via a propensity algorithm in SPSS 28.0 (IBM Corp. Armonk, NY). Descriptive comparisons for matched pairs were performed with paired samples t-test, Wilcoxon signed rank test, and McNemar test depending on distributions (2-sided *p*), with missing data imputed using multiple imputation with chained equations.

## RESULTS

The clinical characteristics at index admission are described in Table 1. The average TNT levels (0.22 ng/ml) were in the normal range for all but case #2 who experienced inferior ST-elevated myocardial infarction confirmed on 12-lead ECG. Four cases presented with elevated creatinine, and sodium levels were all within normal ranges. Most patients presented with pulmonary rales, peripheral oedema, and pulmonary congestion (67-75%), with diuretics the most common intervention (75%). A comparison between groups is presented in Table 2. By comparison to the non-exposed group, we observed differences between groups in New York Heart Association (NHYA) class, diastolic blood pressure (BP), and heart rate (HR) but not systolic BP, saturation of peripheral oxygen (Sp0_2_) or HF phenotypes. There were no differences in TNT, creatinine, sodium, admission use of inotropes, vasodilators or diuretics, readmission, PTSD or other comorbidities between the exposed patients and age-matched female controls (not shown). There was also no difference between cases and controls in hospital readmission at 6 month follow-up.

**Table 1.** Clinical presentation at index heart failure admission of women with a history of physical abuse or sexual abuse or IPV.

|  | **M ± SD or N(%)** | **#1** | **#2** | **#3** | **#4** | **#5** | **#6** | **#7** | **#8** | **#9** | **#10** | **#11** | **#12** |
| --- | --- | --- | --- | --- | --- | --- | --- | --- | --- | --- | --- | --- | --- |
| TNT ng/ml | 0.22 ± 0.58 | 0.02 | 2.06* | 0.02 | 0.02 | 0.04 | 0.02 | 0.27 | 0.09 | 0.02 | 0.08 | 0.02 | 0.02 |
| NT-proBNP | - | 3067 |  |  | 9975 | 16486 |  |  |  |  |  |  |  |
| Creatinine<br>μmol/L | 110.82 ± 75.16 | 74 | 81 | - | 116* | 117* | 81 | 170* | 315* | 61 | 73 | 59 | 72 |
| Sodium mmol/L | 138.18 ± 2.64 | 139 | 136 | - | 139 | 139 | 139 | 134 | 143 | 134 | 139 | 140 | 138 |
| Pulmonary rales | 9 (75) | Yes | No | No | Yes | Yes | Yes | Yes | Yes | Yes | No | Yes | Yes |
| Jugular venous<br>pressure | 6 (50) | No | No | No | Yes | Yes | Yes | Yes | Yes | No | No | Yes | No |
| Peripheral<br>oedema | 8 (66.67) | Yes | No | No | Yes | Yes | Yes | Yes | Yes | Yes | No | No | Yes |
| Hepatomegaly | 0 (0) | No | No | No | No | No | No | No | No | No | No | No | No |
| Peripheral<br>hypoperfusion | 0 (0) | No | No | No | No | No | No | No | No | No | No | No | No |
| Pulmonary<br>congestion | 8 (66.67) | Yes | No | No | Yes | Yes | No | Yes | Yes | Yes | No | Yes | Yes |
| Inotropes | 3 (25) | No | Yes | No | No | No | No | No | No | No | Yes | No | Yes |
| Vasodilators | 0 (0) | No | No | No | No | No | No | No | No | No | No | No | No |
| Diuretics | 9 (75) | Yes | No | No | Yes | Yes | Yes | Yes | Yes | Yes | No | Yes | Yes |

\* above reference values; TNT, Troponin T;

**Table 2.** Comparison of matched pairs on index admission outcome variables.

|  | **HF control** | **HF violence exposure** | **P** |
| --- | --- | --- | --- |
|  | **N = 12** | **N = 12** |  |
| Age yrs | 53.8 ± 14.1 | 55.0 ± 11.3 | 0.40 |
| HF phenotype |  |  | 0.25 |
| HFpEF, (>50%) | 50.0 | 16.7 |  |
| HFmEF (40-49%) | 8.3 | 16.7 |  |
| HFrEF (≤39%) | 41.7 | 66.7 |  |
| HF aetiology |  |  | 0.09 |
| Ischemic | 33.3 | 16.7 |  |
| Hypertensive | 8.3 | - |  |
| Valve disease | - | - |  |
| Arrhythmia | 8.3 | 25.0 |  |
| Cardiomyopathy | 33.3 | 41.7 |  |
| Infective | 16.7 | 16.7 |  |
| NYHA class II | 58.3 | 8.3 | **0.04** |
| III | 41.7 | 83.3 |  |
| IV | - | 8.3 |  |
| Systolic BP mmHg<sup>a</sup> | 104.3 ± 8.7 | 105.3 ± 9.5 | 0.80 |
| Diastolic BP mmHg<sup>a</sup> | 64.1 ± 4.0 | 60.1 ± 3.9 | **<0.01** |
| Heart rate per min<sup>a</sup> | 66.3 ± 3.3 | 55.0 ± 7.3 | **<0.001** |
| Peripheral oxygen saturation (SpO<sub>2</sub>)<sup>a</sup> | 96.0 ± 0.7 | 94.9 ± 1.9 | 0.11 |
| HF readmission within 6 months | 16.7 | 16.7 | 0.99 |
| Post-traumatic stress disorder or | 16.7 | 33.3 | 0.25 |
| adjustment disorder |  |  |  |

*Data presented as % unless otherwise denoted<sup>a</sup> $M \pm SD$ ; EF, ejection fraction; p, preserved; m, mild; r, reduced; HF, heart failure;*

## DISCUSSION

The majority of women with HF exposed to IPV and abuse presented with pulmonary symptoms (rales, congestion) and peripheral oedema, hence, diuretics were the most common intervention. These cases were marked by more severe NYHA class compared to their non-exposed counterparts. The finding of higher NYHA class and lower diastolic BP and HR suggests reduced vagal tone and decreased cardiac output. Prior work in HF implicates different physiological mechanisms underlying BP and HR responses to stress; endothelial function and autonomic cardiac drive respectively [15]. The current findings parallel reports of diminished cardiovascular reactivity to stress in HF [15] and abuse populations [16]. Specifically, HF patients with the lowest BP and HR reactivity to a mental stressor were at higher risk of mortality [15]. Exposure to abuse, especially sexual abuse, was associated with lower systolic BP and HR reactivity [16] leading to the suggestion that blunted cardiovascular reactivity is representative of dysregulation in the hypothalamic-pituitary-adrenal (HPA) axis. Blunted HPA axis response has been observed in female survivors of childhood abuse [17] and parallels recent work showing decreased functioning in the thalamus on fMRI among survivors of childhood sexual abuse [18].

Some epidemiological evidence has suggested early life psychosocial adversity is unrelated to CVD risk factors in adulthood, however, such research included parental death and neglect as psychosocial risk factors in addition to sexual abuse [19]. Generally, however, research consistently reports a preponderance to subclinical CVD risk factors such as hypertension, obesity, diabetes, as well as CVD in survivors of abuse or IPV [2-5]. The current findings did not support prior work [2, 3] and a potential explanation for our discrepant findings is that prior reports have utilized questionnaires covering childhood exposure to abuse, which are subject to memory and recall biases when used in adult populations. Another possible explanation is that all patients were hospitalized with HF and a non-HF control group may show different results. Nonetheless, these findings are submitted with limitations including the small retrospective case-series and inclusion of only female HF patients, thus the findings may not generalize to males and other CVD populations.

In conclusion, our case-series and comparison with age-matched peers raise the possibility that IPV and sexual abuse is associated with blunted cardiovascular reactivity and dysregulated HPA-axis functioning in female patients hospitalised with HF. These findings have implications for trauma informed healthcare including that female HF patients with trauma history may present with blunted cardiovascular reactivity and dysregulated HPA-axis functioning.

## Data Availability

Due to the potential for re-identifying sensitive information about the exposure of interest, only general descriptive information is provided for each case. Data is not available due to ethics restraints.

